# Lung ultrasound findings in patients with novel SARS-CoV2

**DOI:** 10.1101/2020.05.18.20105775

**Authors:** Mark E. Haaksma, Micah L. A. Heldeweg, Jorge E. Lopez Matta, Jasper M. Smit, Jessica D. van Trigt, Jip S. Nooitgedacht, Carlos V. Elzo Kraemer, Armand R.J. Girbes, Leo Heunks, David J. van Westerloo, Pieter R. Tuinman

## Abstract

**Background:** Over 2 million people worldwide have been infected with Severe Acute Respiratory Distress Syndrome Corona Virus 2 (SARS CoV2). Lung ultrasound has been proposed to diagnose and it. However, little is known about ultrasound findings in these patients. Our aim is to present an overview of lung ultrasound characteristics in critically ill patients with SARS CoV2 pneumonia overall and in relation to the duration of symptoms and clinical parameters.

**Methods:** On the Intensive Care Unit of two academic hospitals, adult patients who tested positive for SARS-CoV2 were included. Images were analyzed for pleural line characteristics, number and appearance of B-lines, BLUE-profiles (Bedside Lung Ultrasound in Emergency), pathology in the PLAPS (Postero Lateral Alveolar and Pleural Syndrome) point and a LUS-score (lung ultrasound). The primary outcomes were frequencies, percentages and differences in lung ultrasound findings overall and between short (≤14 days) and long (>14 days) duration of symptoms and their correlation with clinical parameters.

**Results:** In this pilot observational study, 61 patients were included with 75 examinations for analysis. The most prevalent ultrasound findings were decreased lung sliding (36%), thickening of the pleural line (42%) and a C-profile per view (37%). Patients with “long” duration of symptoms presented more frequently with a thickened and irregular pleural line (21% (32) vs 9% (11), p=.01), C-profile per patient (47% (18) vs. 25% (8), p=.01) and pleural effusion (19% (14) vs 5% (3), p=.02) compared to patients with short duration of symptoms. Lung ultrasound findings did not correlate with P/F ratio, fluid balance or dynamic compliance, with the exception of the LUS-score and dynamic compliance (R^2^=0.27, p=.02).

**Conclusion:** SARS CoV2 results in significant ultrasound changes, with decreased lung sliding, thickening of the pleural line and a C-profile being the most observed. With time, a thickened and irregular pleural line, C-profile and pleural effusion become more common findings.

## Introduction

At the time of writing, close to 2,5 million people worldwide have been infected with Severe Acute Respiratory Distress Syndrome Corona Virus 2 (SARS CoV2), of whom approximately 150.000 have died. The disease is quickly spreading, necessitating quick adaptation of clinicians, hospitals and existing protocols. The latter are changing on a daily basis according to novel insights which have been emerging constantly. A lot of discussion has arisen regarding the optimal approach for imaging of infected patients since the gold standard for thoracic imaging, chest computed tomography (CT), poses an additional risk of spread of infection since it necessitates transportation of patients.

Bedside lung ultrasound may very well be the modality of choice, as it has high sensitivity for detecting pathology at the lung surface, such as pleural thickening, consolidation and ground glass like patterns as seen on CT.(1,2) Recent literature also demonstrates that ultrasound outperforms chest X ray (CXR) in detecting these pathologic entities.(3)

This makes lung ultrasound an excellent tool for diagnosing and monitoring of disease progression, as it offers no exposure to radiation, does not require transport and therefore also saves direly needed personal protective material. However, due to the novelty of the disease, there is a scarcity of data related to the typical lung ultrasound findings which may be observed in patients infected with SARS CoV-2. In addition, we do not know if lung ultrasound can be used for monitoring of disease progression, as it is unknown how findings may change throughout the course of the disease and if they correlate to clinically relevant disease related parameters.

We therefore aim to present an outline of lung ultrasound findings in critically ill SARS CoV-2 patients overall, in relation to duration of symptoms, and to determine if there is a correlation between ultrasound findings and physiological parameters such as the P/F-ratio (ratio between partial oxygen pressure and fraction of inspired oxygen).

## Methods

### Study design and population

This study was conducted in two academic intensive care units (ICU’s) (Amsterdam UMC, location VUmc, the Netherlands and LUMC, Leiden, the Netherlands). The protocol to utilize data gathered during routine ultrasound was approved by the local ethics board (Registration ID: 2020.011). The necessity for informed consent was waived. The trial was registered in the Dutch trial registry (Netherlands Trial Register (registration ID: NL8540). Patients were followed up until discharge, death or when still admitted on the ICU until submission of the manuscript to this journal.

The study population consisted of adult (>18 years) ICU patients, who tested positive for SARS-CoV2 at least once before admission. Sex, age, weight, height, days from hospital and ICU admission, time spent on the ventilator before ultrasound examination, SOFA score (Sequential Organ Failure Assessment) on the day of examination, ventilator settings, inflammatory markers, and serum creatinine were recorded. Data were derived form a dedicated patient data management system and data closest to the time of examination were used.

Two groups were defined based on symptom duration from their onset, where ≤ 14 days was defined as “short group” and >14 days as “long group”, which was arbitrarily chosen although based on the clinical observation that the disease often worsens after 10-14 days.

### Ultrasound Measurements

All images were acquired by lung ultrasound certified clinicians, using a Sonsosite-EDGE II or Philips Lumify ultrasound system. Certification entailed a two-day course and thereafter supervision by a physician with extensive ultrasound experience (> 5 years) until sufficient expertise was reached (a minimum of 30 exams).(4) Researchers (MEH, MLH and JLM) performing offline ultrasound analysis were blinded to the patient’s baseline characteristics.

All measurements were made according to the BLUE-protocol, consisting of two ventral-(Upper BLUE-point and Lower BLUE-point) and one dorso-lateral point (PLAPS-point (postero lateral alveolar and pleural syndrome)) point of measurement(s), on either side of the thorax.(5)

Ventral measurements were performed using a 10-5 MHz linear transducer (VUmc) or the Lumify L12-4 linear array transducer which has a 12 to 4 MHz extended operating frequency range (LUMC), both in lung setting and with image depth set at >6 centimeters. This depth was chosen based on the hospital’s local guideline on ultrasound acquisition as to ensure standardization of imaging. (6) The PLAPS measurements were made with a 5-1 MHz cardiac transducer (Amsterdam) of with the Philips S4-1 broadband phased array probe which has 4-1 MHz operating frequency ranger (LUMC), with settings freely adjustable by the operator to obtain an ideal image. In one center (LUMC), in addition to the aforementioned protocol, the Lung Ultrasound (LUS)-score, a 12-region protocol was performed as well.(7) In this center the number of B-lines are not routinely measured due to presumed low reproducibility.(8)

In each image the following analyses were made: 1) Movement of pleural line as present, decreased or absent 2) the pleural line was described as either normal, thickened, irregular or thickened and irregular 3) the total number of B-lines 4) the appearance of B-lines as either <3, ≥3 and separated or ≥3 coalescent 5) the BLUE-profile per view as either A, B or C profile and 6) the BLUE-profile per patient as A (was taken together with A’ due to low number), A/B, B (was taken together with B’ due to low number) or C profile 7) in case of a C-profile, as subpleural or translobar consolidation 8) PLAPS as either absent or present with consolidation and/or pleural effusion. (7)

### Statistical Analysis

No sample size calculation was performed as this study was meant to be a pilot study to give insights into baseline findings in patients with SARS CoV-2. Statistical analyses were performed using SPSS IBM version 22 (SPSS Inc., Chicago, IL, USA). Variables were tested for normality with the Shapiro-Wilk test, evaluation of histograms and Q-Q. plots. Descriptive statistics are presented as means ± standard deviations (±SD), medians and interquartile range [IQR] or numbers (percent %) when appropriate. Differences in characteristics between duration of symptoms/ventilation, the ≤ 14 and >14 days of symptoms groups, were tested with an independent-samples t-test, Mann-Whitney U test or Chi-squared test when appropriate. Correlations for nominal and continuous variables were tested with generalized linear models and with linear regression for continuous variables.

Analyses were made per patient and view, equalling 6 views per patient. The LUS score was calculated based on 6 views per hemithorax but analysed as one data-point per patient.

In one center (LUMC) the total amount of B-lines per view was not counted and perceived as missing data, while in the other center (VUmc) the LUS-score was not calculated and perceived as missing data. Statistical analyses were performed using two-sided hypothesis tests; a p-value of < 0.05 was regarded statistically significant.

## Results

This study was performed from March 27^th^ 2020 until April 20^th^ 2020. Patient enrolment is summarized in figure 1. A total of 93 patients were screened of which 61 patients were included, with in total 75 lung ultrasound examinations and 450 images/views to be analysed. Of these, 2.9% (13) were missing. For the lung ultrasound characteristic divided into short and long duration of symptoms, 5 patients (30 views = 6.7% of all views) were not included due to missing information on the start date of symptoms.

**Figure 1.**
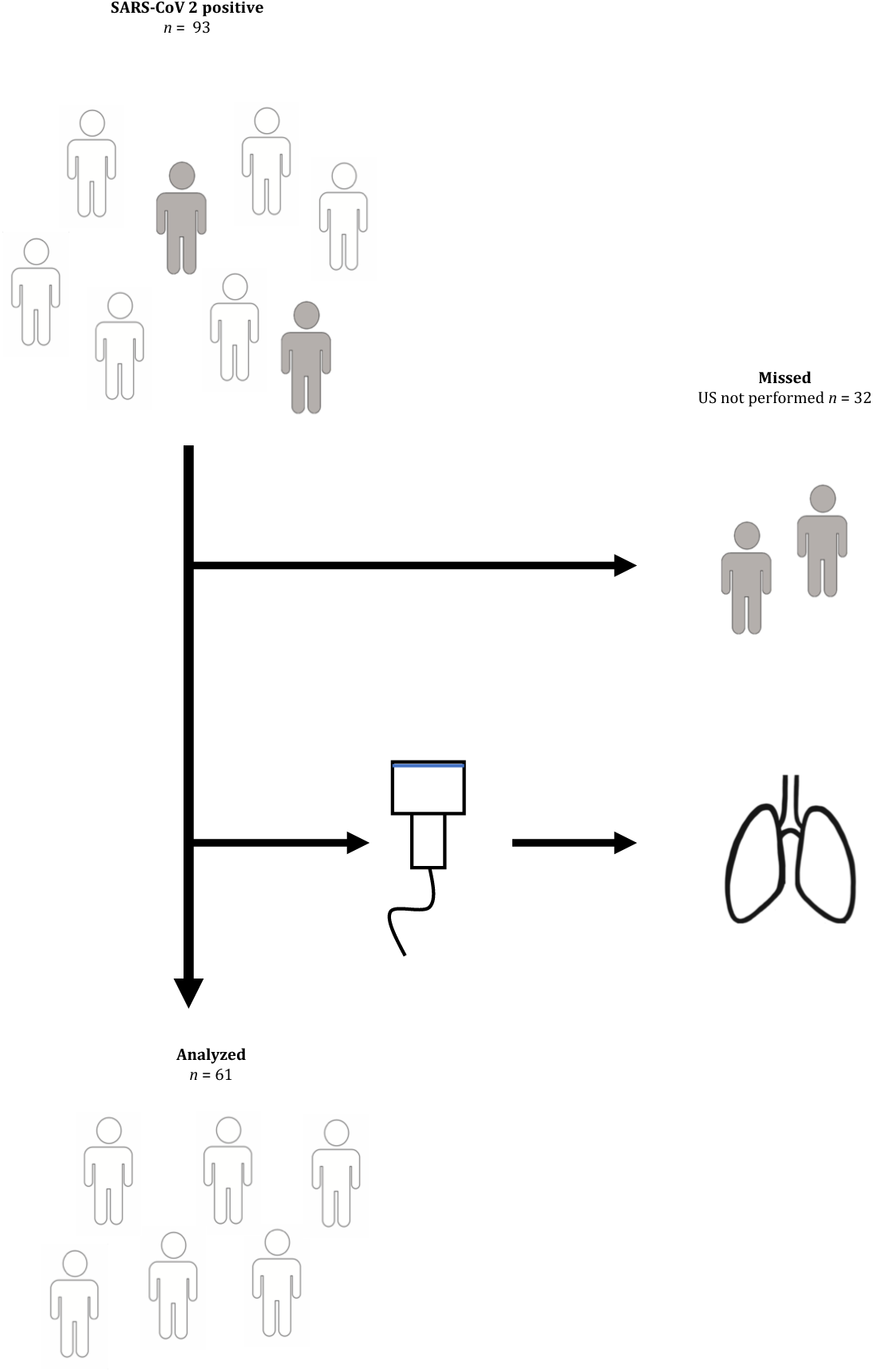
Flowchart of patient inclusion.

### Baseline characteristics

Baseline characteristics are presented in Table 1. The only statistically relevant difference was found in the white blood cell count 8.7 [5.2-12.3] vs. 13.8 [7.8-17.7 (p=.01) for the short and long symptom duration group, respectively. A trend was observed towards higher levels of PEEP in the long symptom duration group. In addition, an overall BMI of 28.3 (±0.7) was found and 90% of the included patients was male. No differences were found in baseline characteristics between the missed and included patients (BMI 28.6 (±0.6) p=.91; Gender, male 81% p=.22 and Age 62 [54-70] p=.10).

**Table 1.**
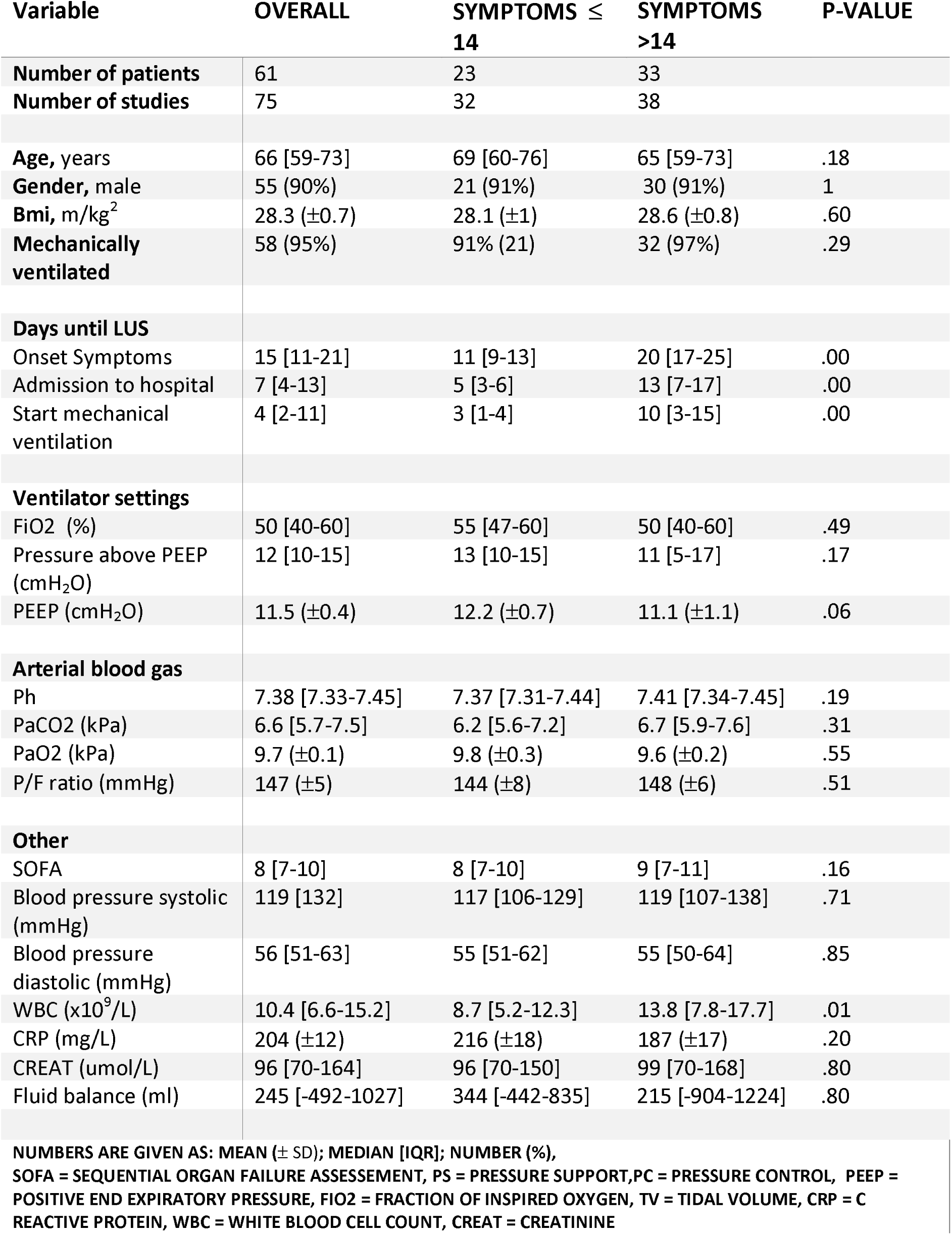
Baseline characteristics of patients included.

| Variable | OVERALL | SYMPTOMS ≤<br>14 | SYMPTOMS<br>>14 | P-VALUE |
| --- | --- | --- | --- | --- |
| **Number of patients** | 61 | 23 | 33 |  |
| **Number of studies** | 75 | 32 | 38 |  |
| **Age, years** | 66 [59-73] | 69 [60-76] | 65 [59-73] | .18 |
| **Gender, male** | 55 (90%) | 21 (91%) | 30 (91%) | 1 |
| **Bmi, m/kg<sup>2</sup>** | 28.3 (±0.7) | 28.1 (±1) | 28.6 (±0.8) | .60 |
| **Mechanically ventilated** | 58 (95%) | 91% (21) | 32 (97%) | .29 |
| **Days until LUS** |  |  |  |  |
| Onset Symptoms | 15 [11-21] | 11 [9-13] | 20 [17-25] | .00 |
| Admission to hospital | 7 [4-13] | 5 [3-6] | 13 [7-17] | .00 |
| Start mechanical ventilation | 4 [2-11] | 3 [1-4] | 10 [3-15] | .00 |
| **Ventilator settings** |  |  |  |  |
| FiO2 (%) | 50 [40-60] | 55 [47-60] | 50 [40-60] | .49 |
| Pressure above PEEP (cmH<sub>2</sub>O) | 12 [10-15] | 13 [10-15] | 11 [5-17] | .17 |
| PEEP (cmH<sub>2</sub>O) | 11.5 (±0.4) | 12.2 (±0.7) | 11.1 (±1.1) | .06 |
| **Arterial blood gas** |  |  |  |  |
| Ph | 7.38 [7.33-7.45] | 7.37 [7.31-7.44] | 7.41 [7.34-7.45] | .19 |
| PaCO2 (kPa) | 6.6 [5.7-7.5] | 6.2 [5.6-7.2] | 6.7 [5.9-7.6] | .31 |
| PaO2 (kPa) | 9.7 (±0.1) | 9.8 (±0.3) | 9.6 (±0.2) | .55 |
| P/F ratio (mmHg) | 147 (±5) | 144 (±8) | 148 (±6) | .51 |
| **Other** |  |  |  |  |
| SOFA | 8 [7-10] | 8 [7-10] | 9 [7-11] | .16 |
| Blood pressure systolic (mmHg) | 119 [132] | 117 [106-129] | 119 [107-138] | .71 |
| Blood pressure diastolic (mmHg) | 56 [51-63] | 55 [51-62] | 55 [50-64] | .85 |
| WBC (x10<sup>9</sup>/L) | 10.4 [6.6-15.2] | 8.7 [5.2-12.3] | 13.8 [7.8-17.7] | .01 |
| CRP (mg/L) | 204 (±12) | 216 (±18) | 187 (±17) | .20 |
| CREAT (umol/L) | 96 [70-164] | 96 [70-150] | 99 [70-168] | .80 |
| Fluid balance (ml) | 245 [-492-1027] | 344 [-442-835] | 215 [-904-1224] | .80 |
| **NUMBERS ARE GIVEN AS: MEAN (± SD); MEDIAN [IQR]; NUMBER (%),**<br>**SOFA = SEQUENTIAL ORGAN FAILURE ASSESSEMENT, PS = PRESSURE SUPPORT, PC = PRESSURE CONTROL, PEEP =**<br>**POSITIVE END EXPIRATORY PRESSURE, FIO2 = FRACTION OF INSPIRED OXYGEN, TV = TIDAL VOLUME, CRP = C**<br>**REACTIVE PROTEIN, WBC = WHITE BLOOD CELL COUNT, CREAT = CREATININE** |  |  |  |  |

### Ultrasound

Ultrasound variables are presented in Table 2 (per view) and Table 3 (per patient). Lung sliding was absent (n=3) or decreased in 36% of views and did not differ between groups. An unaffected pleural line was seen in 36% of views, with a thickened pleural line being seen in 42% of them. A thickened and irregular pleural line was significantly more frequently seen in patients with long vs short symptom duration group, 21% (32) vs. 9% (11), respectively. According to the BLUE-protocol, an A-, A/B, B- and C-profile were seen in, 26%, 23%, 14% and 37% of patients respectively, thus in 60% compatible with pneumonia. The C-profile was seen significantly more frequently in the long symptom duration group 47% (18) vs. 25% (8). B-line types were classified as more than three and separated (26% (79)) or coalescent (13% (38)) and did not differ between groups. Overall, the PLAPS point did not show pathology in 39% of views, and when positive consolidation (46%) was the most frequent finding. A “typical” ultrasound exam is presented in Figure2. A trend was observed towards fewer pathological findings in the short symptom duration group, while pleural effusion was seen more frequently in the long symptom group (5% (3) vs. 19% (14)).

**Table 2.** Lung ultrasound findings in critically ill patients with SARS CoV2 pneumonia per view.

| VARIABLE | OVERALL | SYMPTOMS ≤ 14 | SYMPTOMS >14 | P VALUE |
| --- | --- | --- | --- | --- |
| **LUNGSLIDING** | 304 | 127 | 152 |  |
| PRESENT | 64% (19) | 66% (84) | 61% (92) | .39 |
| DECREASED | 35% (105) | 33% (42) | 39% (59) | .30 |
| ABSENT | 1% (3) | 1% (1) | 1% (1) | 1 |
| **PLEURAL LINE** | 304 | 127 | 152 |  |
| NORMAL | 36% (110) | 44% (56) | 32% (49) | 0.04 |
| THICKENED | 42% (129) | 43% (54) | 42% (64) | .87 |
| IRREGULAR | 5% (16) | 5% (6) | 5% (7) | 1 |
| THICKENED AND IRREGULAR | 16% (48) | 9% (11) | 21% (32) | .01 |
| **BLUE PROFILE PER POINT** | 304 | 127 | 152 |  |
| A | 46% (139) | 56% (71) | 39% (59) | .01 |
| B | 38% (117) | 35% (45) | 41% (62) | .31 |
| C | 15% (47) | 9% (11) | 20% (31) | .01 |
| **NUMBER OF B-LINES** | (n=51) | (n=21) | (n=24) |  |
| TOTAL PER PATIENT | 7 (±0.6) | 7 (±1) | 7 (±0.9) | .85 |
| **B-LINE TYPE** | 302 | 127 | 151 |  |
| ≥ 3 SEPARATED | 26% (79) | 26% (33) | 26% (39) | 1 |
| ≥ 3 COALESCENT | 13% (38) | 10% (13) | 15% (23) | .21 |
| **C PROFILE TYPE** | 48 | 11 | 22 |  |
| SUBPLEURAL | 92% (44) | 91% (10) | 95% (21) | .66 |
| TRANSLOBAR | 8% (4) | 9% (1) | 5% (1) | .66 |
| **PLAPS** | 139 | 56 | 73 |  |
| NO PATHOLOGY | 39% (54) | 45% (25) | 32% (23) | .13 |
| PLEURAL EFFUSION | 12% (17) | 5% (3) | 19% (14) | .02 |
| CONSOLIDATION | 46% (64) | 45% (25) | 48% (35) | .73 |
| PLEURAL EFFUSION AND CONSOLIDATION | 3% (4) | 5% (3) | 1% (1) | .17 |
| **LUNG SCORE** | (n=24) | (n=10) | (n=14) |  |
| TOTAL PER PATIENT | 19,5 (±1) | 18.4 (±1) | 20.4 (±1.5) | .24 |

**Table 3.**
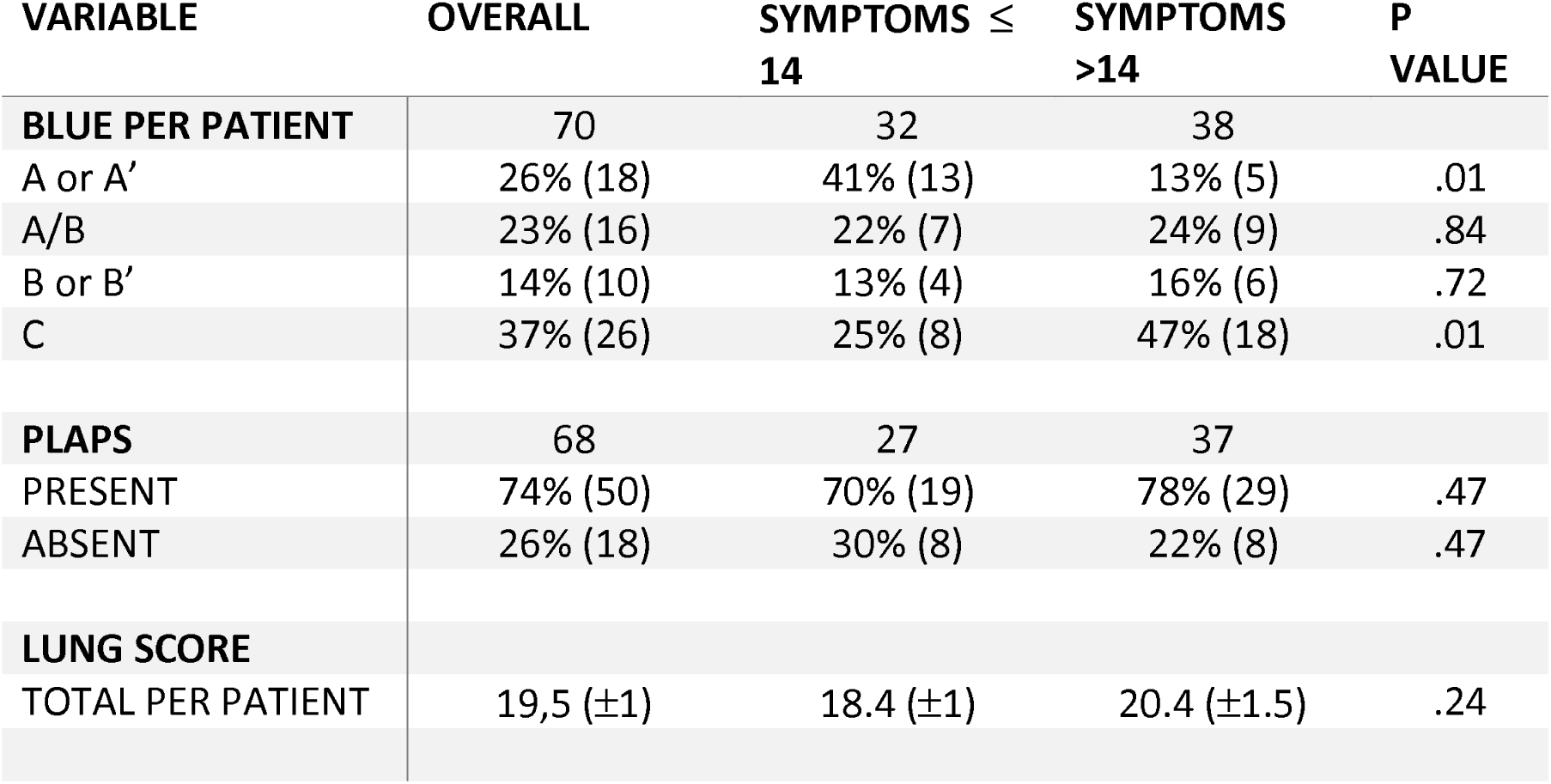
Lung ultrasound findings in critically ill patients with SARS CoV2 pneumonia per patient.

**Figure 2.**
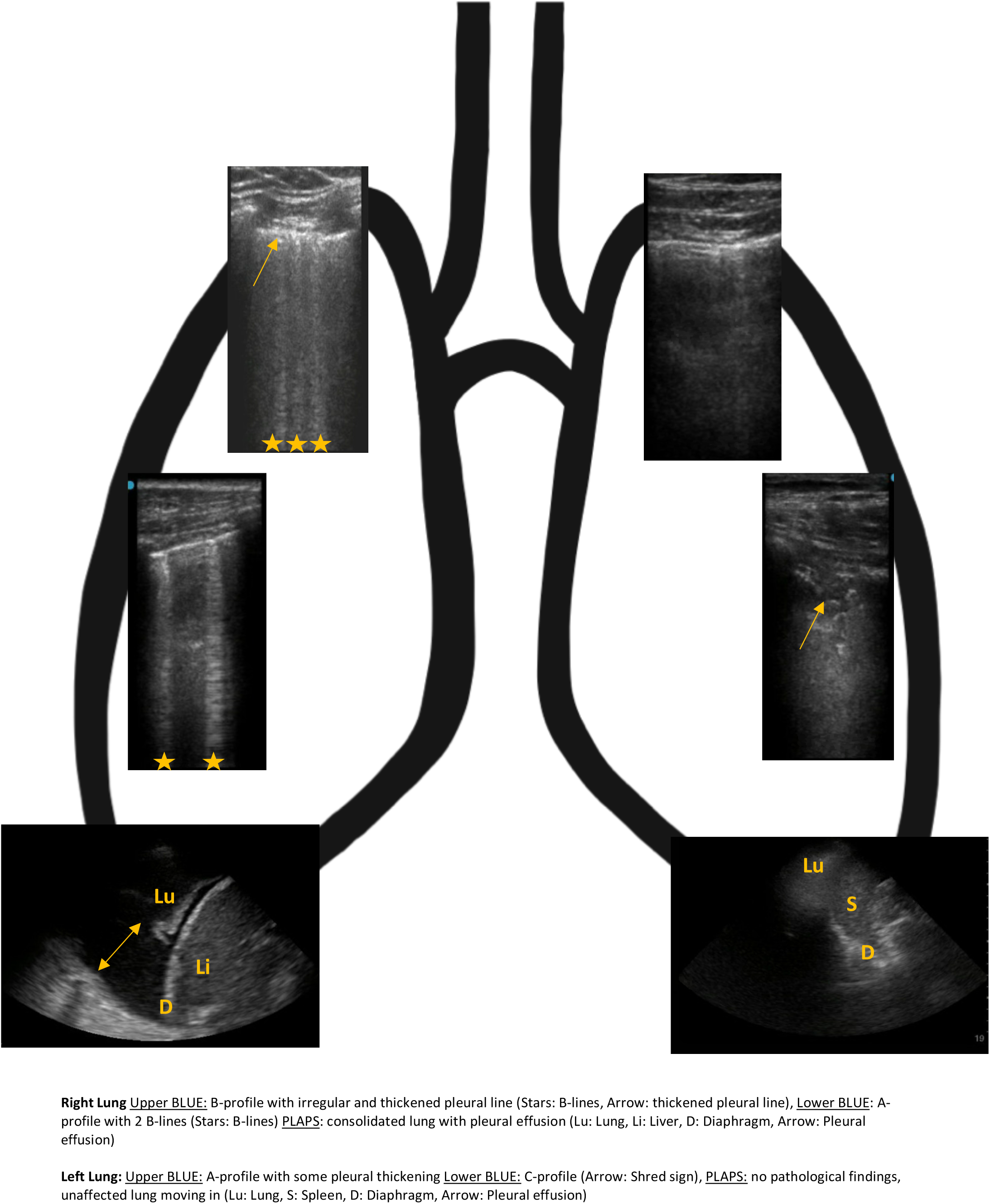
Lung ultrasound findings in patients with SARS CoV2 pneumonia.

A LUS-score was calculated in 24 patients, with a mean of 19.5 (± 1). The score did not differ between symptom duration groups. There was no correlation between BLUE-profile per patient with P/F-ratio (p=.29), fluid balance (p=.84) or dynamic compliance (p=.19). For the LUS-score, a correlation was observed with compliance (p=.02, R^2^=0.27), a trend for fluid balance (p=.09) but not correlation with PF-ratio (p=.98).

## Discussion

The main findings of this study are that 1) SARS CoV2 results in significant ultrasound changes on the lung in most patients, with decreased lung sliding, thickening of the pleural line and a C-profile being the most observed, however with a large part of cases still showing an A-profile. 2) In patients with long duration of symptoms (>14 days) compared to those with a short duration (≤ 14 days), a thickened and irregular pleural line, C-profile and pleural effusion are more common. 3) The BLUE-protocol did not correlate with P/F-ratio, fluid balance or dynamic compliance. 4) The LUS-score is weakly correlated with compliance.

Over the last weeks, several case-reports already described changes of the pulmonary parenchyma due to SARS CoV-2. However, these were only presented in a very small number of patients without baseline characteristics or standardized ultrasound approach.(2,9) To our knowledge, this is the first study that accurately presents a comprehensive overview of ultrasound findings in a large cohort. Our findings are comparable to ARDS, with varying patterns of interstitial syndrome and consolidation, however with a lower rate of pleural effusion.(10,11)

Over time, the occurrence of these findings change, with an increase of a thickened and irregular pleural line, C-profile and pleural effusion. This is in line with previous articles that demonstrated comparable changes on CT.(12-14) With this in mind, we think that lung ultrasound poses a valuable alternative for monitoring disease progression in the context of this pandemic.(15) Especially considering that it does not require transport and therefore not only saves direly needed personal protective equipment but also limits the necessity to take patients out of isolation. However, it is crucial to realize, that 26% of all cases were found to have an A-profile, indicating a non-pathological state of the lungs when assessing the BLUE-points. This comes not completely as a surprise, given that on CT the lung parenchyma is very heterogeneously affected and pathological regions might be missed. At this point, we wish to mention, that in all but one case with A-profile, either thickening of the pleural line, present PLAPS or one BLUE-point showing a B-line pattern was seen. It is therefore an important consideration when lung ultrasound examinations are performed, especially when used as a diagnostic modality in patients with suspected SARS CoV-2 infection, to select a more comprehensive approach such as the 12-region protocol or to also interpret pleural line thickening or a local B-line pattern indicative of disease.(7) We hypothesize that this becomes especially relevant in patients presenting to emergency ward, as in this population abnormalities in the PLAPS point or pleural thickening might be less frequently encountered, thus leaving an import part of patients with a negative lung ultrasound examination.

We also set out to determine whether the BLUE-profile was correlated to currently relevant parameters such as the P/F-ratio, fluid balance or dynamic lung compliance, which was not the case. We hypothesize that this might be attributed to several factors. While the 12-region protocol obviously covers a larger area of the lung than the BLUE-protocol, a large part of the most dorsal regions is not visualized and therefore not accounted for. In addition, even if extensive, lung ultrasound only examines the outermost parts of the pulmonary parenchyma, while studies show that on CT also deeper lung parts are affected.(12-14) This might be especially relevant for the “H”-type, and less for the “L”-type lungs, as latter present mostly with subpleural-, and the former also with deeper regions affected.(16) Also, while an “L”-type patient might have a high LUS-score due to extensive subpleural groundglass pattern (on ultrasound as B-line pattern), they still might have near normal compliance. Nevertheless, a weak, but statistically significant correlation was found between the LUS-score and dynamic compliance. This should merely be seen as a hypothesis generating finding and larger studies with more standardized measurements with static compliance and are needed to test it.

## Strengths

The strength of this study is its size with an overview of lung ultrasound findings in SARS-CoV2 positive patients and reporting both on the use of the BLUE-protocol and LUS-score. This contributes greatly to the currently available body of evidence. In addition, the study was carried out in two different hospitals, by multiple operators and using two different ultrasound approaches, thereby increasing its external validity.

## Limitations

This study has several important limitations. Firstly, the sample was based on availability of ultrasound. Especially in the first days of the outbreak with a large number of admissions some patients did not receive and ultrasound examination. With the introduction of better logistics and dedicated proning-teams this changed and we were able to examine every admitted patient. Secondly, the length of symptom duration for subgroup analysis was arbitrarily chosen. In addition, we were not able to correlate our findings to endpoints such as mortality or extubation outcome, as the majority of the patients is still admitted and ventilated at the time of writing.

## Conclusion

SARS CoV2 pneumonia results in significant ultrasound changes, with decreased lung sliding, thickening of the pleural line and a C-profile being the most observed. With time, a thickened and irregular pleural line, C-profile and pleural effusion become more prevalent. Lung ultrasound seems to be a valuable alternative for CT in diagnosing and monitoring SARS CoV-2 pneumonia.

## Data Availability

The data will be made available for other researchers on reasonable request, upon publication in an indexed journal.

## Guarantor statement

Mark Evert Haaksma takes responsibility for the content of the manuscript, including the data and analysis.

## Authors’ contributions

MHa, MHe, AG, LH, CK, JM, DW and PT were responsible for the conception and design of the work. MHa, MHe, JN, JT, JM, PT were responsible for acquisition and or analysis of the data. MHa, MHe, JM were responsible for building the database. MHa and PT were responsible for drafting the manuscript and all authors provided critical revisions for it. All authors read and approved the final manuscript and ensured that questions related to the accuracy or integrity of any part of the work were investigated and resolved.

## Ethics approval and consent to participate

Approval was given by the local ethics committee (METc (Medisch Ethische Toetsings Commissie) VUmc) with the study number 2020.11. Consent for participation was not applicable as ultrasound measurements were carried out as part of routine clinical examination.

## Consent for publication

Consent for publication was waived by the local ethics board.

## Availability of data and materials

The datasets used and/or analyzed during the current study are available from the corresponding author on reasonable request.

## Abbreviations list

Creat: Creatinine
BLUE: Bedside Lung Ultrasound in Emergency
CI: Confidence Interval
CoV: Corona Virus
COVID: Corona Virus Disease
CRP: C Reactive Protein
CT: (chest) Computed Tomography
CXR: Chest X-ray
FiO2: Fraction of Inspired Oxygen
IQR: Inter Quartile Range
LUS: Lung Ultrasound
METC: Medisch Ethische Toetsings Commissie (Medical Ethics Board)
PaO2: Partial Oxygen Pressure
PEEP: Positive End Expiratory Pressure
PLAPS: Postero Lateral Alveolar and Pleural Syndrome
PC: Pressure Control
PS: Pressure Support
P/F ratio: Ratio between Partial Oxygen Pressure and Fraction of Inspired Oxygen
SARS: Severe Acute Respiratory Distress Syndrome
SD: Standard Deviation
SOFA: Sequential Organ Failure Assessment
WBC: White Blood cell Count

